# Disentangling physiological heterogeneity in retinal aging using a deep learning–based biological age framework

**DOI:** 10.64898/2026.02.13.26346265

**Authors:** Renxiang Chu, Aimin Sun, Jinfeng Qu, Meng Lu

## Abstract

Biological age estimators quantify aging-related variation but provide limited insight into organ-specific aging processes. The retina enables non-invasive visualization of microvascular and neural structures and has emerged as a promising modality for biological age prediction. However, existing retinal aging models typically produce unidimensional age estimates with limited interpretability. Here we develop a deep learning framework based on a large-scale vision foundation model to estimate retinal biological age from fundus images and to characterize the physiological heterogeneity underlying retinal aging. Using a reference cohort of 56,019 relatively healthy individuals, the model achieved a Mean Absolute Error of 2.48 years in age prediction. Analysis of age deviations in a real-world clinical cohort (n = 46,369) revealed non-linear associations with cardiometabolic risk and population heterogeneity in aging patterns. Integrating multidimensional physiological profiling, feature attribution and unsupervised analysis, we identified distinct retinal aging signatures associated with systemic inflammation and hemodynamic variation. To further characterize age-related deviations, we introduced a residual learning framework that decomposes retinal aging signals into a normative age-related component and additional components associated with physiological variation, achieving a Mean Absolute Error of 1.80 years on the independent healthy test set. This approach provides an interpretable representation of retinal aging and a framework for studying organ-level aging processes and their relationship to systemic health using large-scale imaging data.

## Introduction

Biological aging constitutes the predominant risk factor for chronic diseases and mortality^1^. However, chronological age alone fails to capture the profound heterogeneity in health status observed among age-matched individuals^1,2^. In recent years, the advent of “Aging Clocks”—estimators of biological age derived from molecular omics or imaging traits—enables the precise quantification of accelerated or decelerated aging^3,4^. Ranging from DNA methylation clocks and routine clinical biochemical indices to plasma proteomic classifiers, these tools have demonstrated remarkable potential in predicting mortality risk and disease susceptibility^2,3,5–7^. Nevertheless, existing molecular biomarkers are often constrained by invasive sampling requirements, temporal fluctuations, and an inability to resolve organ-specific aging trajectories^2,8^. Crucially, systemic clocks may obscure the spatial heterogeneity of aging, where pathological alterations in specific organs often precede systemic decline^9,10^. Consequently, there is an urgent need to develop non-invasive, organ-specific imaging biomarkers capable of capturing these distinct biological aging processes to facilitate both aging research and clinical translation.

The retina serves as the unique anatomical window allowing for direct, non-invasive visualization of the microvasculature and central nervous system tissue, where vascular morphology and neurodegenerative changes profoundly reflect systemic cardiovascular and metabolic health status^11,12^. In recent years, deep learning models have achieved breakthroughs in age prediction from ophthalmic imaging, narrowing the prediction error to within 2–3 years and establishing robust associations with cardiovascular mortality, stroke, and systemic metabolic risks^13–15^. However, these models remain inherently unidimensional scalar predictors: they predominantly oversimplify the aging process into a single metric (i.e., Δage), failing to elucidate which specific physiological systems drive these deviations or to distinguish between healthy rejuvenation and pathological masking. This lack of interpretability poses a substantial risk of clinical misinterpretation — while existing studies adopt the default assumption that younger is better, they overlook the possibility that certain pathological states (e.g., vascular remodeling) may alter tissue morphology to manifest a “pseudo-youth” phenotype^16^. Furthermore, current models largely assume that the drivers of aging remain constant throughout the lifespan, thereby neglecting the temporal heterogeneity of the aging process.

Aging is not merely a linear progression but is characterized by distinct “phase transitions” at specific chronological milestones. Recent multi-omics evidence reveals that molecular aging undergoes profound non-linear shifts at approximately 44 and 60 years of age^17^, paralleling findings that brain network instability reaches a critical inflection point during midlife^18^. This implies that the drivers of accelerated aging are likely stage-dependent, with fundamental distinctions between early-life accelerators (e.g., acute inflammatory storms) and late-life contributors (e.g., chronic metabolic accumulation)^7^. However, existing “one-size-fits-all” models trained across the entire lifespan fail to capture these temporal dynamics.

Spatially, systemic aging emerges from the interplay of hemodynamic, metabolic, immune, and renal systems, yet these systems age asynchronously^9^. Recent insights into pathological masking suggest that certain pathological states or interventions may induce a facade of functional restoration by suppressing inflammatory phenotypes, thereby maintaining a macroscopically young appearance despite underlying cellular senescence^16^. Based on this, we hypothesize that extreme negative Δage values do not necessarily indicate superior health but may instead mask pathological remodeling, such as that induced by hypertension. Consequently, there is a critical need to pivot from unidimensional scalar prediction to multidimensional structural deconstruction. Such an approach is essential to resolve spatiotemporal heterogeneity and elucidate the specific physiological dimensions driving retinal aging across different life stages.

Recently, medical foundation models—empowered by self-supervised pretraining on massive, heterogeneous datasets—have demonstrated robust feature extraction capabilities that surpass traditional supervised learning, catalyzing a transition in medical AI from task-specific models to generalist representation learning^19^. In ophthalmology, vision foundation models pretrained on millions of retinal images—exemplified by Vision Transformer-based Masked Autoencoders (MAE)^20^ — have achieved expert-level performance in disease screening tasks^21,22^. However, current models predominantly focus on discrete diagnostic classification. Their inherent ‘black-box’ nature precludes the deconstruction of the intrinsic multidimensional structure of continuous physiological variables, such as aging. Furthermore, the prevalence of shortcut learning poses significant challenges, undermining both AI fairness and generalizability^23,24^. Consequently, harnessing the potent representation capabilities of foundation models to bridge the gap from unidimensional prediction to multidimensional construct validation remains a central challenge in contemporary medical AI.

To address these challenges, we leverage VisionFM^21^—a large-scale vision foundation model pretrained on millions of multimodal ophthalmic images—to establish a state-of-the-art retinal biological age-prediction benchmark, achieving an unprecedented Mean Absolute Error (MAE) of 2.48 years, surpassing previous benchmarks which typically range from 2.86 to 3.55 years for CNNs^15,25^, and edging out recent transformer-based models (MAE ∼2.50 years)^26^. Transcending traditional end-to-end prediction, we integrated multidimensional physiological profiling with deep feature attribution and unsupervised clustering. This approach confirmed the nonlinear spatiotemporal heterogeneity of retinal aging and unveiled the dynamic evolution of key drivers across the aging spectrum. Specifically, it enabled the stratification of individuals into distinct phenotypic subgroups, identifying that while systemic inflammation drives accelerated aging, specific hemodynamic anomalies characterize a “pseudo-youth” profile. Crucially, to resolve the pathological masking effect and validate these drivers, we developed a disentangled physiological residual learning framework. By employing orthogonal constraints, we decomposed high-dimensional signals into “Base” aging (representing normative senescence) and pathological “Bias” (driven by specific physiological systems). This decoupled model significantly outperformed benchmarks in healthy populations, a performance gain that reciprocally validates the statistical independence and additivity of the identified physiological drivers. Quantitatively, the disentangled framework achieved a MAE of 1.80 years on the independent healthy test set, a significant improvement over the baseline (2.48 years) that validates the precision of our normative aging definition. Consequently, this work transforms the retinal clock from a “black-box” tool into an interpretable monitoring system, providing a theoretical foundation to deconstruct nonlinear aging mechanisms and guide personalized anti-aging interventions.

## Results

### Construction of a Deep Learning-based retinal aging clock

In the research, we developed an age prediction model based on pre-trained foundation model VisionFM^21^ using a Vision Transformer (ViT)-based Masked Autoencoder (MAE) in a self-supervised manner, which was trained on 3.4 million ophthalmic images integrating multi-source imaging devices and modalities. Foundation models acquire robust feature representations by training on large-scale imagery, offering significant potential for deep information mining. To construct the aging clock, we employed the VisionFM backbone by freezing the pre-trained weights to extract high-dimensional deep features from input fundus images. These features were then fed into a trainable Multi-Layer Perceptron (MLP) regression head, which was optimized using chronological age as the ground-truth label (Fig. 1A).

**Fig. 1.**
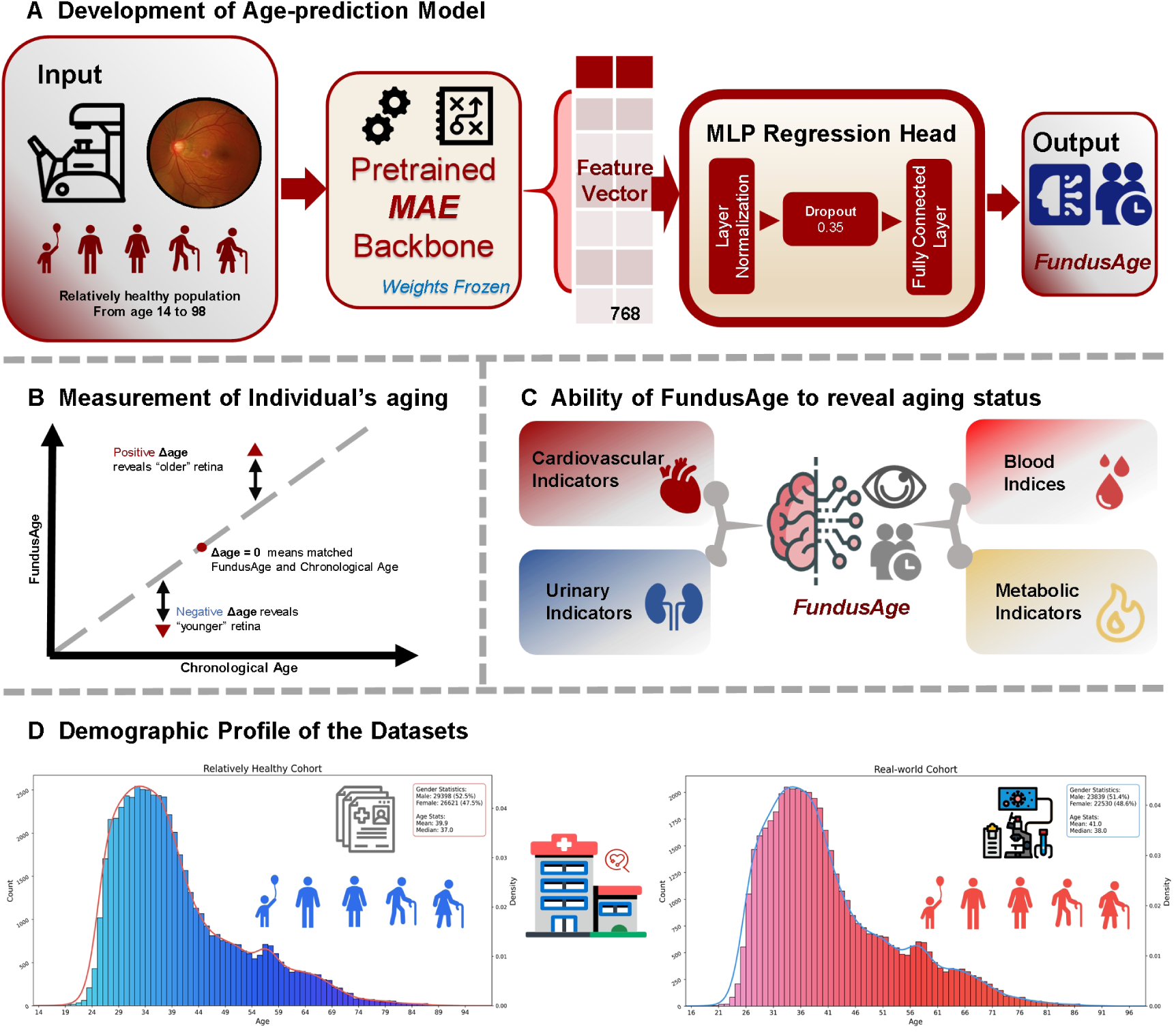
Overview of the research workflow. (A) We developed a deep learning model to predict biological age from retinal fundus photographs, using a dataset of 112,019 total images (from left and/or right eyes, derived from ∼50,000 relatively healthy individuals) which was split into training (N=89,615), validation (N=11,202), and test (N=11,202) sets. The model architecture employs a pretrained Masked Autoencoder (MAE) backbone to extract features, which are then processed by an MLP regression head. (B) The model predicts an individual’s FundusAge, from which we define Δage as the difference between the predicted FundusAge and the individual’s chronological age. (C) Based on the established model, we analyzed the relationship between fundus age and various physiological and biochemical indicators from multiple dimensions. (D) Demographic profiles of the two cohorts included in this study.

To establish a robust baseline for normal physiological aging, we curated a reference dataset of 56,019 relatively healthy participants (aged 14 to 98 years) who had no reported history of systemic diseases. This cohort, contributing a total of 112,019 fundus photographs, served as the standard for “healthy aging.” Detailed baseline characteristics of this cohort are provided in Supplement Table 1. As visualized in Fig. 1D, the demographic profile exhibited a relatively balanced sex ratio and a natural, right-skewed age distribution, effectively capturing the representative long-tail structure of the general population.

To ensure rigorous evaluation, the healthy dataset was randomly partitioned into a training set (N=89,615 images), a validation set (N=11,202), and an independent test set (N=11,202). The model predicts an individual’s FundusAge, and the deviation from chronological age is defined as Δage (FundusAge minus Chronological Age). This metric serves not only as an indicator of accelerated or decelerated retinal aging (Fig. 1B) but also as a pivotal systemic biomarker. We utilized Δage to dissect the intricate associations between retinal aging and a comprehensive array of somatic health dimensions, spanning cardiovascular, metabolic, and biochemical indicators, thereby establishing a link between ocular phenotypes and systemic physiological status (Fig. 1C).

### Performance evaluation of FundusAge

To rigorously evaluate the model’s performance and generalization capability, we assessed it on the independent test set. The model demonstrated exceptional predictive accuracy, characterized by a strong linear alignment between predicted FundusAge and chronological age (Fig. 2A), achieving a Pearson correlation coefficient of 0.9613 (R^2^ = 0.9219) between predicted FundusAge and chronological age, with a Mean Absolute Error (MAE) of only 2.48 years. Further examination of prediction errors to rule out systematic bias revealed that the distribution followed a zero-centered Gaussian distribution of prediction errors (Fig. 2B), and that Bland-Altman analysis confirmed high agreement with a negligible mean bias of +0.16 years and narrow 95% limits of agreement (-6.21 to +6.53 years) (Fig. 2D). Crucially, regression analysis of Δage against chronological age revealed a near-flat slope of -0.033 (Fig. 2C). This suggests that our re-weighting and balanced sampling strategies effectively mitigated the ‘regression to the mean’ effect, ensuring consistent accuracy across the entire age spectrum by preventing the systematic overestimation of young and underestimation of elderly individuals.

**Fig. 2.**
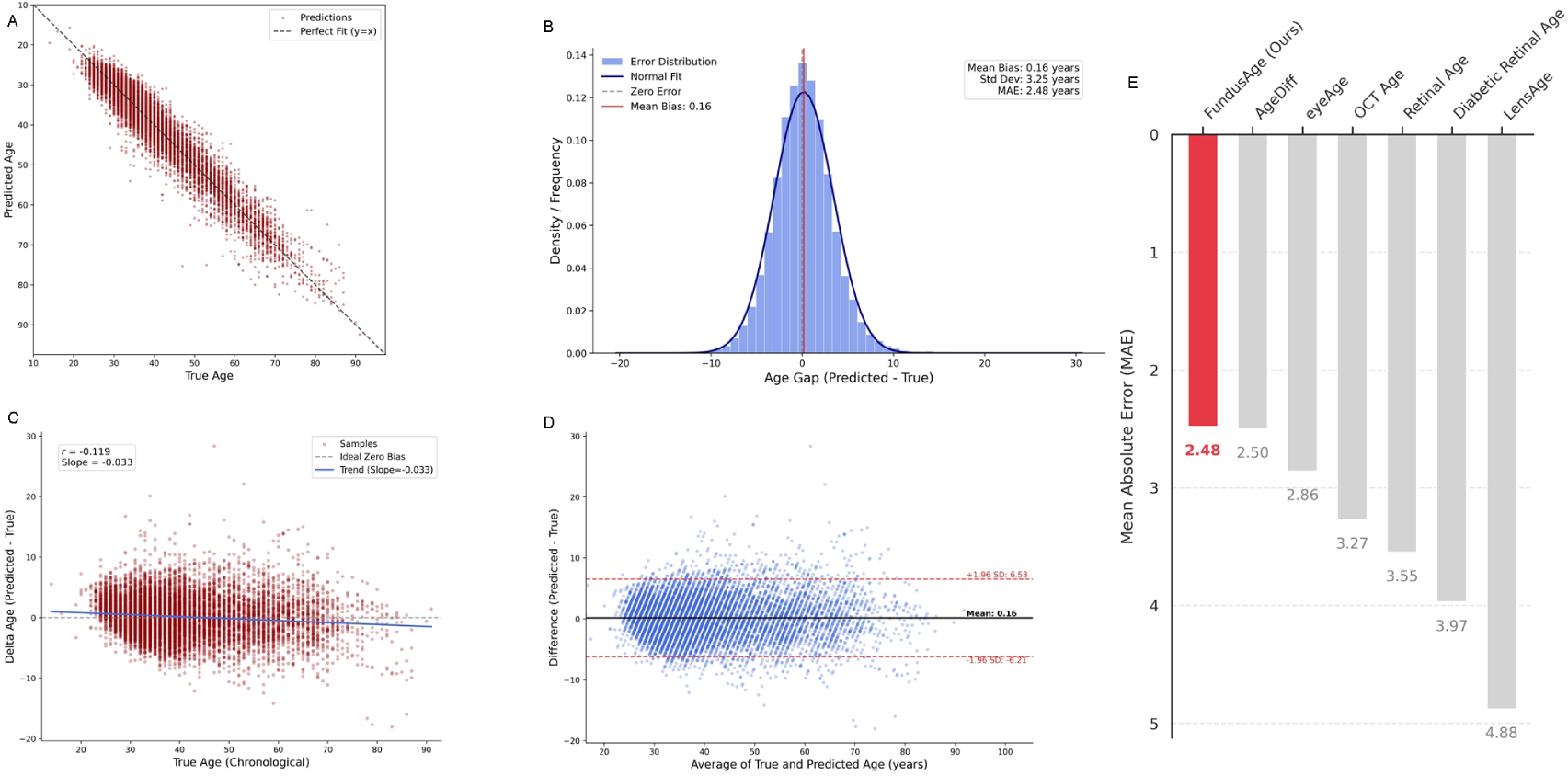
Analysis and evaluation of the deep learning model’s prediction performance on the independent test set (N=11,202). (A) Scatter plot of predicted age versus chronological age in the test cohort. (B) Histogram of the age delta distribution in the test cohort. The age delta follows a normal distribution, with a mean of only +0.16 years, indicating no systematic over- or under-estimation bias. (C) Scatter plot of Δ age vs. chronological age. The near-zero slope (-0.033) and low correlation (r = -0.119) indicate no significant age-dependent bias in the predictions. (D) Bland-Altman plot for agreement analysis between predicted and chronological age in the test cohort. The 95% limits of agreement (LoA) were between -6.21 and +6.53 years, demonstrating the model’s reliability. (E) Performance comparison of retinal age estimation models. FundusAge outperforms existing state-of-the-art methods, achieving the lowest Mean Absolute Error (MAE) of 2.48 years.

FundusAge achieves state-of-the-art performance across all compared modalities and architectures. Specifically, our model achieved the lowest reported MAE and the highest goodness of fit, outperforming models based on other modalities (e.g., OCT, Lens) and traditional CNNs. Notably, it also surpasses recent Transformer-based approaches (e.g., AgeDiff), demonstrating the superior feature mining potential and scalability of the MAE-ViT foundation model. Beyond accuracy, the results uniquely highlight the clinical utility of our marker, establishing its statistical independence from routine biochemical indices and revealing distinct underlying aging mechanisms, rather than solely associating with mortality risks.

### Bidirectional Deviations Signal Systemic Health Risks and Dynamic Disease Burden

Leveraging our constructed Deep Learning model, we applied the FundusAge model to a cohort of 46,369 participants recruited from real-world clinical departments in a hospital setting (demographic and feature distributions are detailed in Figure 1D and Supplement Table 1). This comprehensive dataset integrates binocular fundus images with age and 30-dimensional physiological and biochemical profiles, allowing us to derive the FundusAge and Δage for each individual.

To systematically explore the clinical significance of the deviation between chronological and biological age, we stratified the population into eight octiles (Q1–Q8) based on Δ age. Notably, the relationship between Δ age and key cardiovascular and metabolic risk indicators (including systolic blood pressure, LDL-C, fasting blood glucose, etc.) did not follow a simple linear trajectory; instead, a distinctive non-linear, U-shaped association was evident (Supplementary Figure 1). Participants in the intermediate octiles (Q2–Q6), where Δ age was close to zero, exhibited the lowest risk levels, representing a homeostatic “healthy baseline.” In contrast, individuals at both extremes of the spectrum—those with significantly accelerated aging (Q7–Q8) and those with a ‘decelerated aging’ phenotype (Q1)—demonstrated elevated levels of somatic health risks. This finding challenges the initial interpretation of Δ age, suggesting that while a positive value indicates accelerated aging, an extreme negative value does not solely reflect a “younger” phenotype but may also signal deviations from physiological homeostasis.

To further elucidate the pathological determinants behind these deviations, we analyzed retinal aging trajectories across different chronic disease states (Fig. 3). The analysis revealed that systemic metabolic and vascular diseases contribute to the distinct divergence in retinal aging patterns. Crucially, in the age range of 20 to 70 years, patients across all disease cohorts consistently exhibited a positive Δ age compared to healthy controls. This indicates that the cumulative physiological burden imposed by chronic conditions is visibly reflected in the fundus as a generalized acceleration of biological aging. However, a paradoxical crossover effect was observed in the advanced age groups (>75 years), where disease trajectories converged with or dipped below the healthy baseline (Fig. 3). While this phenomenon may partly reflect survival bias—where individuals with the most accelerated aging phenotypes experience earlier mortality, leaving a resilient survivor cohort—it unmistakably highlights that FundusAge is sensitive to both the accumulation of pathological stress in mid-life and the shifting survival landscapes in late life.

**Fig. 3.**
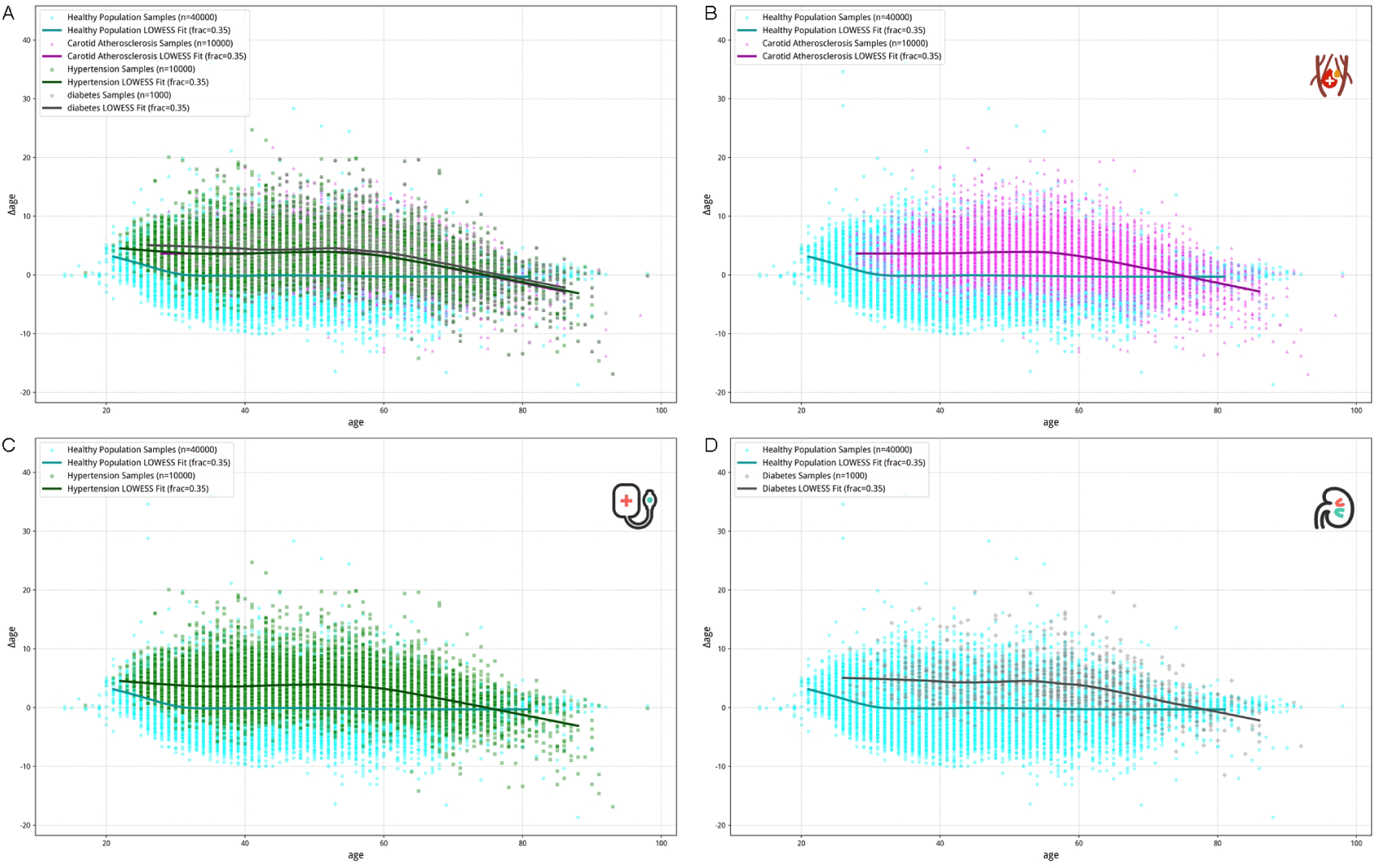
Distribution and LOWESS-fitted curves of Δage against chronological age for healthy individuals and individuals with systemic diseases. In the healthy control cohort, the LOWESS-fitted curve remains stable around a Δage of 0. In contrast, cohorts of individuals with the three systemic diseases (carotid atherosclerosis, hypertension, and diabetes) all exhibit a positive Δage before approximately 75 years of age and a negative Δage thereafter. (A) Distribution and fitted curves for healthy individuals versus all three systemic disease groups. (B) Comparison between healthy individuals and individuals with carotid atherosclerosis. (C) Comparison between healthy individuals and individuals with hypertension. (D) Comparison between healthy individuals and individuals with diabetes.

### Biological Interpretability and Phenotypic Stratification of FundusAge Divergent physiological basis of aging trajectories

To elucidate the specific biological mechanisms underlying divergent aging trajectories at a granular level, we first employed XGBoost, a supervised learning algorithm with high efficiency in capturing complex non-linear interactions. This approach was utilized to classify participants into decelerated, normal, and accelerated aging phenotypes based on their Δage status. To demystify the model’s decision-making process, we integrated SHAP (Shapley Additive Explanations), a game-theoretic approach that unifies feature importance by calculating the marginal contribution of each physiological feature to the model’s output (Supplementary Figure 2). The analysis revealed a binary differentiation in physiological drivers: Systemic inflammatory indicators were identified as the dominant features for the accelerated aging group (Q7-Q8), suggesting they are the key biological signatures characterizing the retinal phenotype’s shift towards an ‘older’ state. Conversely, the decelerated aging group (Q1-Q2) was dominated by hemodynamic-related indicators, implying that specific vascular morphological changes might confound aging signatures, leading to predictions associated with a pseudo-young phenotype.

### Orthogonality to routine biochemical profiles

Furthermore, to determine whether FundusAge provides biological information beyond traditional physiological and biochemical indicators, we constructed a Tabular Transformer^27^. Unlike traditional linear methods, this architecture leverages self-attention mechanisms to reconstruct global dependency networks within a high-dimensional space, capturing complex non-linear interactions between retinal features and the 30-dimensional physiological profile (Supplementary Figure 3). Visual inspection of the attention weights revealed that while the model accurately reproduced known strong couplings among blood indices (e.g., between hemoglobin and hematocrit), it demonstrated pronounced attention sparsity between retinal aging features and routine markers. This distribution confirms that the aging information encapsulated in fundus imagery is statistically orthogonal to routine laboratory panels, indicating that FundusAge is not a simple redundancy of known markers but serves as an independent, complementary biological dimension. This result underscores that retinal imaging, as a high-dimensional modality, contains rich biological content extending beyond routine clinical indicators.

### Unsupervised Clustering Reveals Population Heterogeneity and Evolution of Aging Patterns

To validate the objectivity of physiological patterns without reliance on a priori labels and to further dissect population heterogeneity, we performed UMAP dimensionality reduction and K-means clustering (k=18) on the high-dimensional feature space using physiological and biochemical data from the real-world cohort (Fig. 4A). The clustering results demonstrated high statistical robustness (Silhouette Score = 0.3793), indicating that the population is not randomly distributed but naturally organized into distinct pathophysiological states (Fig. 4D). Notably, while the UMAP projection colored by Δage shows a continuous gradient, Δage magnitude alone is insufficient to fully resolve the granular biological specificity within the population (Fig. 4B, C). Consequently, we shifted to a refined analysis at the subgroup level. by mapping each subgroup onto a “Chronological Age vs. Δage “ coordinate system (Fig. 4E). Within this landscape, the Homeostatic Baseline (Quadrant Q3) represents a healthy reference state, where clusters like C9 and C12 exhibit physiological indicators within normal ranges or benign deviations. Analyzing the key deviations further elucidated the age-dependent evolution of accelerated aging: Early-onset aging in young individuals (Quadrant Q2) is primarily driven by acute systemic inflammation, exemplified by Cluster C11’s drastic elevations in neutrophils (+41%) and WBCs; conversely, cumulative aging in older populations (Quadrant Q1) is defined by chronic metabolic syndrome, with Cluster C1 displaying severe lipid disorders (+202% Triglycerides) and obesity as core contributors. Most notably, Quadrant Q4 highlighted a critical “Pathological Masking” effect: despite substantial disease burdens, subgroups with severe diabetes (Cluster C14) or hypertension (Cluster C10) paradoxically maintained suppressed Δage levels. Paradoxically, hypertensive features like vascular tortuosity and increased reflectivity create a complex visual texture that mimics the vascular abundance of a youthful retina, leading the model to underestimate biological age despite arteriolar narrowing.

**Fig. 4.**
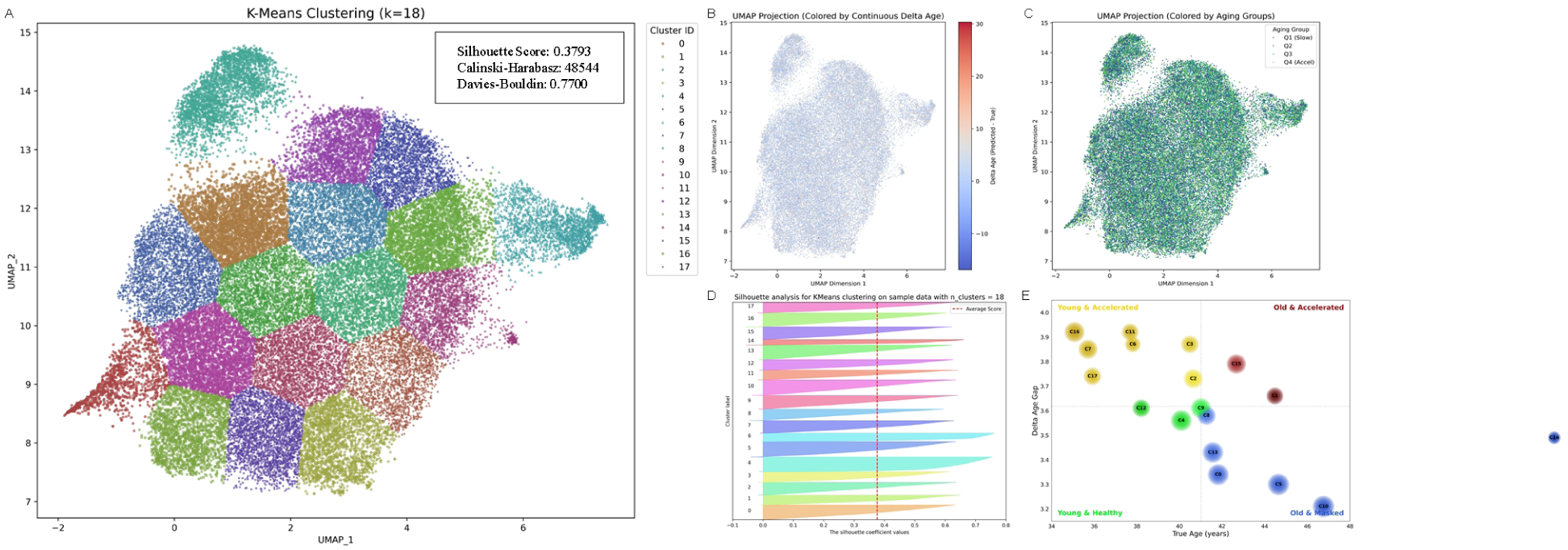
Unsupervised clustering reveals heterogeneity in retinal aging phenotypes within the real-world cohort. We employed Uniform Manifold Approximation and Projection (UMAP) for dimensionality reduction and K-means clustering (k=18) to dissect the health and aging status revealed by Δ age. (A) Distribution of K-means clusters. The inset box presents three key validation metrics: the Silhouette Score (0.3793), measuring cluster cohesion and separation; the Calinski-Harabasz Index (48544), evaluating the variance ratio; and the Davies-Bouldin Index (0.7700), assessing cluster overlap. Collectively, these metrics confirm the robust statistical structure of the identified phenotypes. (B) UMAP projection colored by continuous Δ age values, highlighting the gradient of aging across the population structure. (C) UMAP projection annotated by Δ age quartiles (Q1-Q4), illustrating the spatial segregation of groups with varying aging rates. (D) Silhouette analysis for the 18 clusters. The silhouette coefficients for individual clusters generally surpass the global average score (red dashed line), indicating high intra-cluster cohesion and inter-cluster separation, thereby validating the reliability of the stratification. (E) Bubble plot distribution of sub-clusters based on chronological age versus Δ age. The bubble size corresponds to the population size (N) of each cluster, while the four quadrants delineate distinct aging patterns.

### Physiologically Disentangled Learning and Biological Specificity Verification

To quantitatively dissect the independent contributions of distinct physiological systems to retinal aging and transcend simple correlation analyses, we constructed a Physiologically Disentangled Residual Learning Framework based on a “Teacher-Student” paradigm (Fig. 5A). In this architecture, the Teacher network utilizes the frozen weights of our pre-trained FundusAge model to provide a robust normative aging baseline unaffected by specific pathological noises. The Student network is explicitly designed to learn the pathological deviations from this baseline through a residual learning strategy. By modeling the age gap as an additive residual, we mathematically force the student network to capture only the features responsible for the deviation, leaving the healthy baseline intact. Uniquely, the Student network incorporates five parallel projection heads, each supervised by specific categories of clinical indicators—Hemodynamic, Metabolic, Renal, Hematologic, and Immune. This design forces the high-dimensional retinal features to decouple into orthogonal subspaces, mathematically ensuring that the final predicted biological age is a composite sum of the normative baseline and the distinct biases contributed by each physiological system.

**Fig. 5.**
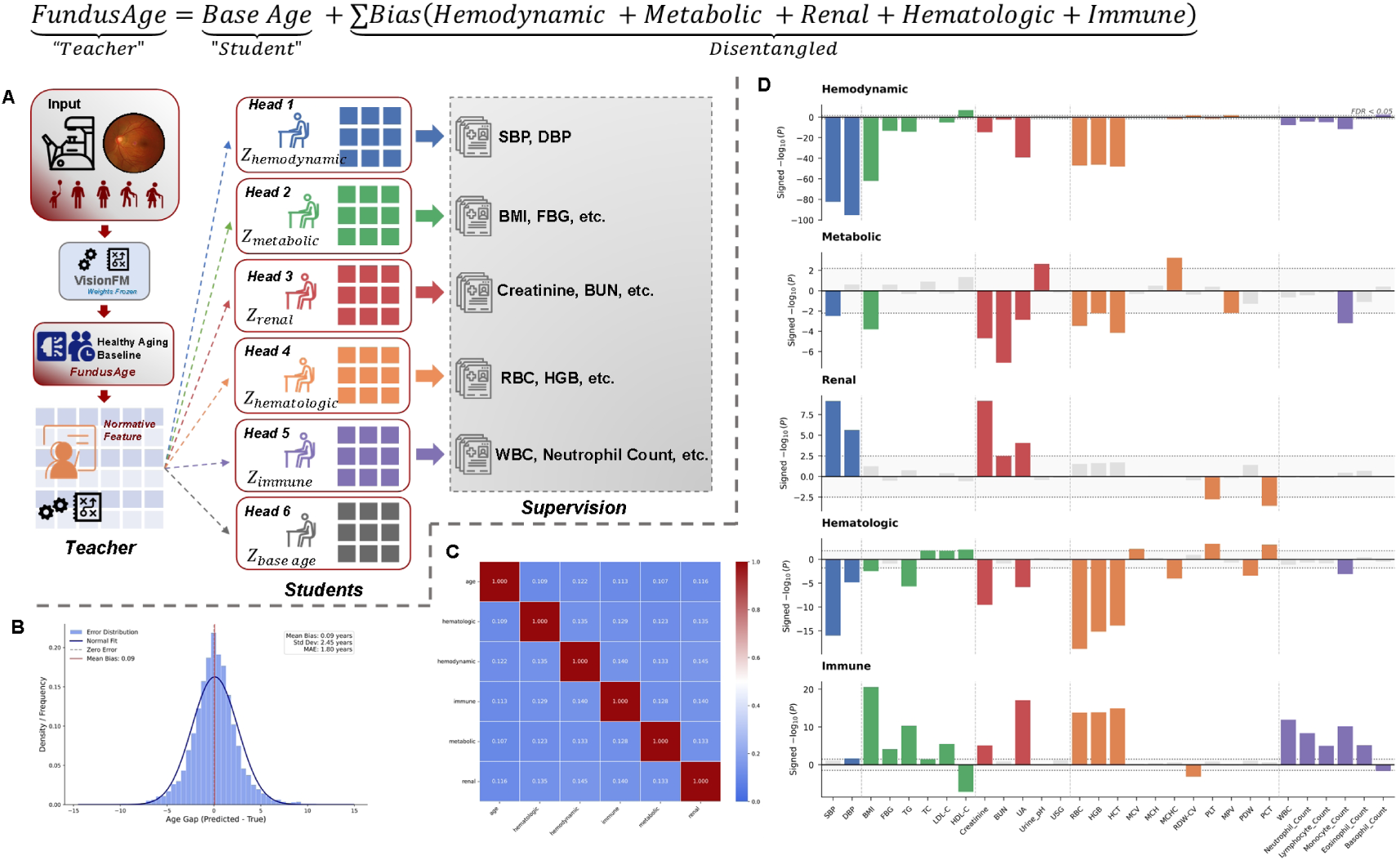
Architecture and Validation of the Physiologically Disentangled Residual Learning Framework. (A) Schematic overview of the proposed framework. A “Teacher-Student” residual learning strategy is employed to decouple physiological aging from pathological interference. The Teacher network (utilizing frozen FundusAge weights) provides a normative aging baseline. The Student network, trained on a mixed-population real-world dataset, incorporates five disentangled heads supervised by grouped clinical indicators. These projection heads map high-dimensional features into orthogonal subspaces to learn specific physiological biases, which are additively combined with the baseline age to refine the final prediction. (B) Performance validation on an independent healthy cohort. The disentangled model exhibits a zero-centered Gaussian error distribution, with a Mean Absolute Error (MAE) significantly superior to that of the baseline model alone, validating the efficacy of the additive correction logic. (C) Orthogonality verification. The Pearson correlation heatmap among the five learned subspaces reveals minimal correlation in off-diagonal regions, demonstrating the successful independent disentanglement of distinct physiological dimensions. (D) Biological specificity analysis. Bar plots display the association between each disentangled subspace and 30 clinical biomarkers. The Y-axis represents the signed − log_10_, reflecting both the direction and statistical significance of the associations. Bars extending up or down indicate positive or negative correlations, respectively. The dashed line indicates the statistical significance threshold adjusted for multiple comparisons (FDR < 0.05). The observation of significant peaks surpassing this threshold within each category confirms that each disentangled head specifically captures its intended biological dimension while effectively filtering out interference from irrelevant indicators.

The implementation of this disentangled architecture yielded superior predictive stability and interpretability compared to the baseline model. When evaluated on the independent healthy test set, the prediction errors of the disentangled model followed a strictly zero-centered Gaussian distribution (Fig. 5B), with a further reduction in Mean Absolute Error to 1.80 years, confirming the validity of the additive residual correction strategy in characterizing healthy aging. Crucially, to verify whether the model successfully separated the physiological signals as intended, we analyzed the correlation matrix between the learned feature subspaces (Fig. 5C). The resulting heatmap exhibited a high degree of diagonality with near-zero off-diagonal correlations, providing strong empirical evidence that the model has effectively disentangled the complex retinal aging signal into mutually exclusive vectors. This orthogonality ensures that the morphological features learned for one dimension, such as vascular mechanical stress, remain statistically independent from others, such as hematological status, thereby avoiding feature redundancy and cross-contamination.

Finally, a Targeted Phenome-Wide Association Study was performed to validate the specific biological semantics captured by each disentangled subspace (Fig. 5D). The Hemodynamic, Renal, and Hematologic subspaces exhibited precise alignment with their supervisory labels, demonstrating high biological specificity: the Hemodynamic head showed exclusive, strong negative associations with Systolic and Diastolic Blood Pressure, confirming its role in capturing vascular filling patterns. Crucially, the Hemodynamic subspace showed a strong negative association with blood pressure, providing mechanistic support for the hypothesis. It implies that the model interprets hypertensive vascular phenotypes—characterized by high tone and tortuosity rather than simple atrophy—as a signal of structural robustness relative to the distinctive vascular dropout seen in healthy aging. This misinterpretation manifests specifically as a negative age bias (pseudo-youth). The Renal head specifically targeted nitrogenous waste products like Creatinine and BUN, while the Hematologic head was strictly associated with red blood cell indices. However, a noteworthy divergence from a priori expectations was observed in the Metabolic and Immune subspaces. Contrary to the assumption that the Metabolic head would capture all biochemical metabolic markers, it primarily exhibited sensitivity to anthropometric features such as Body Mass Index (BMI), while also displaying balanced associations with renal dimension indicators. This pattern likely reflects the impact of structural load and highlights the complex inter-dependencies between physiological information. Intriguingly, the Immune subspace captured a rich array of signals: beyond the expected inflammatory markers (WBC, Neutrophils), it significantly encoded multiple biochemical metabolic indicators, including glucose metabolism, Uric Acid, and Lipid profiles. This observation suggests that within the retinal microenvironment, the morphological manifestations of biochemical metabolic dysregulation are phenotypically inextricably linked with chronic inflammation, corroborating the biological essence of “immunometabolism.” Rather than representing a model error, this reflects the biological reality where metabolic dysregulation and chronic inflammation are inseparable. Consequently, the model successfully learned to attribute these biochemically-driven vascular alterations to the Immune subspace, while reserving the Metabolic subspace primarily for characterizing structural alterations associated with body mass and composite physiological states.

## Discussion

Biological aging is increasingly quantified using data-driven clocks^1,6^; however, most existing approaches implicitly assume that aging is a unidimensional process in which younger biological age uniformly reflects better health^7,15^. In this study, by disentangling retinal aging into base aging and bias aging, we demonstrate that apparent biological age instead emerges from the interaction of two parallel yet intercrossed aging processes within the retinal vascular system.

Base aging captures a stable, monotonic trajectory of senescence that is tightly coupled to chronological time and largely invariant to disease status or physiological conditions^6,8,28^. This component may reflect a intrinsic biological program that governs vascular and neurovascular aging, including progressive loss of microvascular integrity^29^, reduced vascular compliance^30,31^, and decline in neurovascular coupling^32^. The robustness and monotonicity of base aging across individuals suggest that it represents a conserved, time-determined axis of aging—functioning as a fundamental coordinate within the aging manifold that is resistant to external modulation^1^.

In contrast, bias aging from our computational analysis, may reflect condition-dependent physiological remodeling shaped by extrinsic and contextual factors such as cardiometabolic status^31^, inflammatory milieu^16,33^, hemodynamic load^30^, and disease-associated vascular adaptation^31^. Unlike base aging, bias aging exhibits substantial inter-individual heterogeneity and is not constrained to increase monotonically with time^6,34^. Our unsupervised clustering (Fig. 4) explicitly resolved this heterogeneity, mapping distinct pathophysiological states—from acute inflammatory phenotypes in youth to chronic metabolic accumulation in the elderly—that remain obscured in scalar predictions. This component captures both beneficial and deleterious deviations from normative aging trajectories, encompassing the effects of exceptional health, compensatory remodeling, and pathological processes. As such, bias aging defines the extent to which aging can be phenotypically slowed, masked, or partially reversed, while simultaneously explaining why younger predicted biological age may paradoxically associate with elevated disease risk in certain populations.

Importantly, this decomposition provides a mechanistic explanation for the phenomenon of pathological pseudo-youth^15^, whereby disease-driven vascular remodeling produces a superficially youthful retinal phenotype despite ongoing intrinsic senescence^29^. Prior retinal aging studies have reported such paradoxical associations without a unifying biological interpretation^15,35^. Our framework resolves this contradiction by showing that apparent rejuvenation can arise from reductions in bias aging that obscure, rather than alter, the underlying base aging trajectory.

Beyond static interpretation, the disentangled framework reveals that the relative contribution of bias aging is stage-dependent across the lifespan. In earlier adulthood, bias aging appears more strongly influenced by inflammatory and acute metabolic perturbations, whereas in later life it is increasingly dominated by chronic vascular and metabolic remodeling. Strikingly, the Immune subspace spontaneously encoded metabolic indicators (Fig. 5D), computationally mirroring the biological immunometabolism axis and confirming that microvascular inflammation is inextricable from metabolic dysregulation. These findings align with emerging multi-omics and neuroimaging evidence that human aging is characterized by nonlinear transitions and shifting system dominance rather than a single continuous process, and extend such observations to the level of retinal vascular biology^1,7,36^.

Conceptually, base aging corresponds to intrinsic aging—biological processes that are programmed, conserved, and governed primarily by time^1,2^. Epigenomic clocks provide a canonical example of such intrinsic aging at the molecular level^6,8,34^. Here, we demonstrate the existence of a corresponding intrinsic aging axis within the retinal vascular system, indicating that programmed senescence is not confined to molecular layers but is also embedded in organ-level vascular structure. Together with molecular, proteomic, and organ-level aging signatures reported in other systems, our findings support a hierarchical view of human aging in which intrinsic aging manifests consistently across biological scales, while extrinsic modulation introduces variability around this core trajectory.

The distinction between base and bias aging may also have important implications for aging research and clinical translation. While intrinsic aging defines an irreducible component of biological senescence, bias aging may indicate theoretical and practical limits of aging modulation. Interventions may meaningfully alter bias aging—thereby improving healthspan and reducing disease risk—without necessarily modifying the underlying intrinsic aging program. This distinction cautions against overinterpreting apparent biological rejuvenation and emphasizes the need for mechanistically informed aging metrics when evaluating anti-aging strategies.

Methodologically, the use of a large-scale vision foundation model was critical for uncovering this structure. By learning rich, high-dimensional representations from diverse retinal phenotypes, the model enabled separation of subtle but biologically meaningful aging components that would likely be collapsed into a single estimate by conventional end-to-end predictors. This highlights the value of representation learning frameworks not merely for improving prediction accuracy, but for revealing latent biological structure within complex medical data^19^.

Several limitations warrant consideration. First, the predominantly cross-sectional design constrains causal inference regarding individual aging trajectories. Second, while bias aging is strongly associated with systemic physiological states, direct attribution to specific molecular pathways remains indirect. Longitudinal studies and multimodal integration will be essential to determine the reversibility and mechanistic underpinnings of bias aging. Nevertheless, these limitations do not diminish the central finding that retinal aging comprises both intrinsic and condition-dependent components.

In summary, this work provides an interpretable and disentangled framework for retinal aging prediction that moves beyond unidimensional black-box models. By separating time-associated aging signals from condition-dependent variation, our approach enables characterization of retinal aging heterogeneity and offers a framework for investigating pathological masking and population-level aging patterns.

**Table 1:**

| Model Name | Source | Modality | Architecture | Performance | Clinical Utility/Notes |
| --- | --- | --- | --- | --- | --- |
| Retinal Age | Zhu et al. (2023) | Fundus | VGG-19<br>(Classic CNN) | MAE: 3.55 years<br>Pearson r: 0.81 | **Mortality:** Each 1-year increase in Retinal Age Gap (RAG) is associated with a **2% increase** in mortality risk.<br>**Disease:** Associated with Parkinson's (10%), kidney failure (10%), and stroke (4%). |
| eyeAge | Ahadi et al. (2023) | Fundus | ResNet-50<br>(Standard CNN) | MAE: 2.86 years (EyePacs)<br>MAE: 3.30 (UKB)<br>Pearson r: 0.95 | **Longitudinal:** Correctly ordered two visits from the same patient with **71% accuracy**.<br>**Genetics:** Identified distinct genetic loci compared to phenotypic age (PhenoAge). |
| RetiAGE | Nusinovici et al. (2022) | Fundus | Xception<br>(Efficient CNN) | AUROC: 0.968 (Internal)<br>AUROC: 0.756 (External) | **Method:** A classification model predicting if an individual is ≥65 years old (Probability score).<br>**Limitation:** Performance dropped on external UK Biobank data, suggesting ethnic biases. |
| AgeDiff | Wang et al. (2024) | Fundus | Swin Transformer<br>(Transformer-based) | MAE: 2.50 years | **Comparison:** In this study, using retinal images alone yielded an MAE of 2.5 years. |
| Diabetic Retinal Age | Abreu-Gonzalez et al. (2023) | Fundus | CNN-based | MAE: 3.97 years | **Population:** Trained specifically on patients with **diabetes**.<br>**Utility:** RAG was significantly higher in patients with diabetic retinopathy (1.905 years) vs without (0.609 years). |
| LensAge | Li et al. (2023) | Lens Photo<br>(Slit-lamp / Smartphone) | ResNet-50<br>(Standard CNN) | MAE: 4.88 years<br>R<sup>2</sup> score: 0.89 | **Accessibility:** Can be **smartphone-based**, increasing accessibility for self-monitoring.<br>**Risk:** A 1-year increase in LensAge is associated with an **8% increase** in all-cause mortality. |
| OCT Age | Chen et al. (2024) | OCT<br>(B-scans) | ResNet-based<br>(DNN) | MAE: 3.27 years | **Comparison:** Improved over the same team's fundus-based model (MAE 3.55).<br>**Mortality:** A 5-year increase in OCT age gap is associated with an 8% increase in mortality. |
| FundusAge | This Study | Fundus | MAE-ViT<br>(Transformer-based) | MAE 2.48 years<br>RMSE: 3.3127<br>Pearson r: 0.9613<br>R<sup>2</sup> Score: 0.9219 | **Mechanisms:** Unveiled differential drivers (Inflammation vs. Hemodynamics) for accelerated/decelerated aging.<br>**Independence:** Proven orthogonal to 30 routine clinical/biochemical markers via masked attention analysis.<br>**Stratification:** Revealed U-shaped risk associations and survival bias patterns in chronic diseases. |

**Table 2:**

| Quadrant2 | Cluster ID | N | True Age (mean) | Δage (mean) | Top Key Deviations (vs. Global Mean) | Proposed Physiological Phenotype |
| --- | --- | --- | --- | --- | --- | --- |
| Q1: Old & Accelerated (High-Risk Aging) | C15 | 2704 | 42.7 | +3.79 | TG (+51%)<br>Eosinophil (+42%) | Mixed High-Risk (Hyperlipidemia + Inflammation) |
|  | C1 | 2076 | 44.5 | +3.66 | TG (+202%)<br>HDL-C (-20%)<br>BMI (+15%) | Severe Metabolic Syndrome (Obesity) |
| Q2: Young & Accelerated (Early-Onset Aging) | C11 | 2075 | 37.7 | +3.92 (Max) | Neutrophil (+41%)<br>WBC (+30%) | Acute Systemic Inflammation |
|  | C16 | 2939 | 35.1 | +3.92 | TG (-41%)<br>Creatinine (-20%) | Malnutrition / Frailty (Low Metabolic Rate) |
|  | C3 | 2270 | 40.5 | +3.87 | Eosinophil (+135%)<br>Neutrophil (+52%) | Severe Immune Activation (Allergy/Autoimmune) |
|  | C6 | 1812 | 37.8 | +3.87 | TG (-25%)<br>PCT (+24%) | Anemia / Infection Risk |
|  | C7 | 2655 | 35.7 | +3.85 | TG (-46%)<br>Eosinophil (-33%) | Hypolipidemia / Low Immunity |
|  | C17 | 2272 | 35.9 | +3.74 | Basophil (+106%)<br>PLT (+26%) | Atypical Hematological Response |
|  | C2 | 2698 | 40.6 | +3.73 | LDL-C (+15%)<br>TG (-14%) | Pure Hypercholesterolemia |
| Q3: Baseline / Low Risk (Relative Healthy) | C9 | 2908 | 41 | +3.61 | Eosinophil (+20%)<br>Monocyte (+15%) | Baseline Reference Group (Typical Patient) |
|  | C12 | 2239 | 38.2 | +3.61 | TG (-43%)<br>HDL-C (+28%) | Cardio-Protective Profile |
|  | C4 | 3140 | 40.1 | +3.56 | Urine pH (+19%) | Dietary/Alkaline Subgroup |
| Q4: Old & Masked (Pathological Masking) | C8 | 2466 | 41.3 | +3.58 | TG (-25%)<br>Eosinophil (-22%) | Low-Risk / Mild deviations |
|  | C14 | 1181 | 57.6 | +3.49 | FBG (+71%)<br>TG (+57%) | Severe Diabetes (Vasodilation Masking) |
|  | C13 | 3081 | 41.6 | +3.43 | Basophil (-20%)<br>Creatinine (+13%) | Active Renal/Muscle Function |
|  | C0 | 3146 | 41.8 | +3.34 | Basophil (-40%)<br>TG (-29%) | Low-Inflammatory State (Quiescent Immune) |
|  | C5 | 3281 | 44.6 | +3.3 | PLT (-25%)<br>Creatinine (+18%) | High Muscle Mass / Hypermetabolism |
|  | C10 | 3334 | 46.8 | +3.21 (Min) | LDL-C (+25%)<br>DBP (+17%) | Hypertension / Hypercholesterolemia (Vascular Filling) |

## Data Availability

All data referred to in this study are available upon reasonable request to the corresponding author, subject to approval by the Institutional Review Board of Peking University People's Hospital and compliance with relevant data protection regulations. Due to the sensitive nature of clinical patient data and privacy protection requirements, individual-level data cannot be made publicly available.

---

**Supplement Table 1.**

| Baseline Characteristics | Total |  |
| --- | --- | --- |
| No. of participants | 56019 | 46369 |
| Age in years (mean ± SD) | 39.87±11.87 | 41.03±11.99 |
| Distribution of Chronological age, n |  |  |
| <20 | 24 | 11 |
| ≥20 and <30 | 10106 | 6662 |
| ≥30 and <40 | 23911 | 19463 |
| ≥40 and <50 | 10884 | 10107 |
| ≥50 and <60 | 6312 | 5652 |
| ≥60 and <70 | 3562 | 3238 |
| ≥70 and <80 | 979 | 1025 |
| ≥80 | 241 | 211 |
| Sex, n (%) |  |  |
| Female | 26621(47.05%) | 23839(51.4%) |
| Male | 29398(52.95%) | 22503(48.6%) |
| Images |  |  |
| Fundus photographs | 112019 | 92738 |

**Supplement Figure 1.**
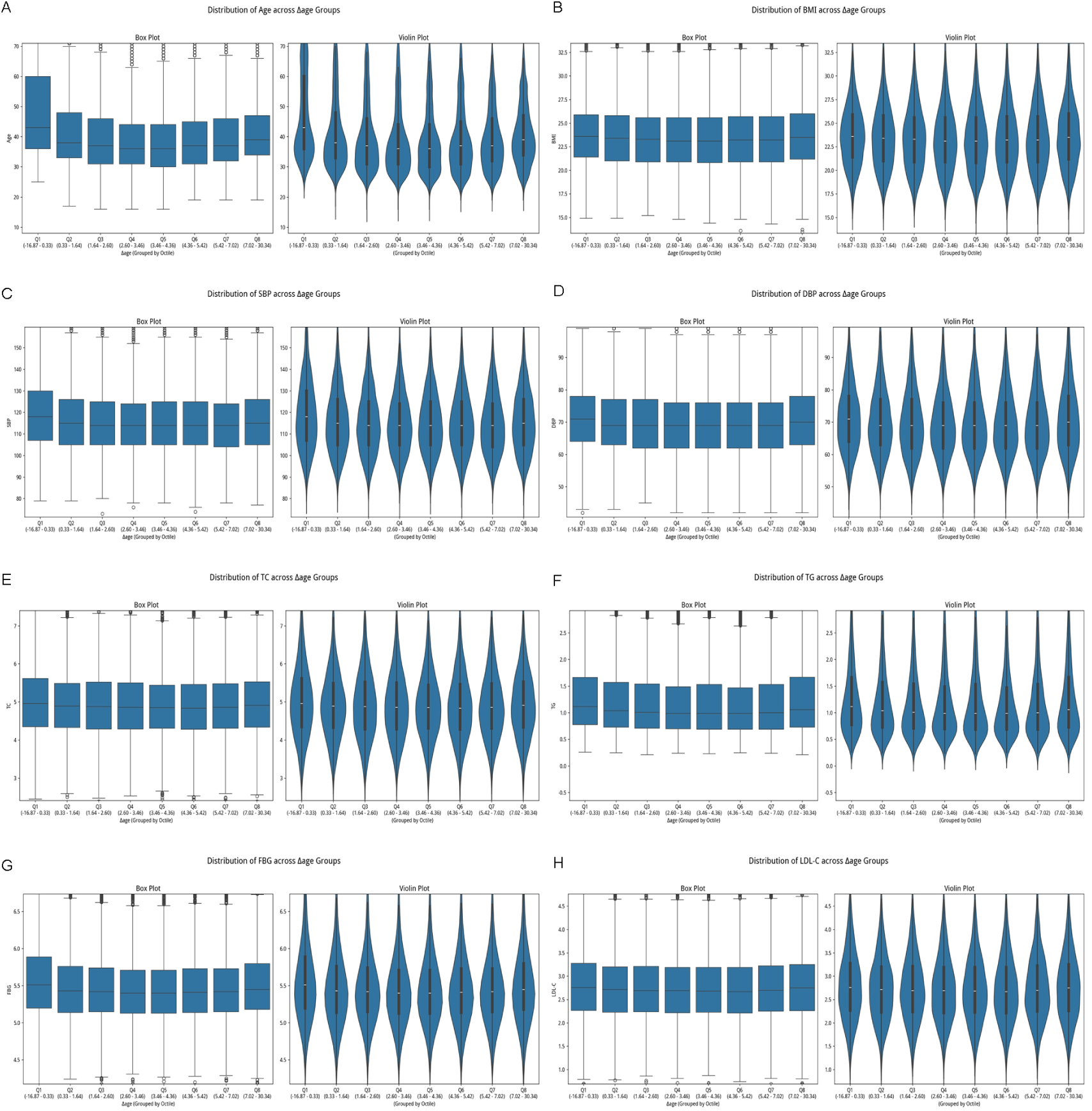
Distribution analysis of Δage with various physiological and biochemical indicators in the mixed-population cohort. We stratified the cohort into eight equal groups (octiles) based on their Δage, from lowest (Q1) to highest (Q8). Boxplots and violin plots were generated for selected indicators within each octile. We observed a distinct U-shaped relationship between Δage and most health risk indicators, suggesting that both extremely low and high Δage values may be associated with accelerated aging or deteriorating health conditions. (A) Association between Δage and chronological age. (B) Association between Δage and BMI (Body Mass Index). (C) Association between Δage and systolic blood pressure (SBP). (D) Association between Δage and diastolic blood pressure (DBP). (E) Association between Δ age and total cholesterol (TC). (F) Association between Δage and triglycerides (TG). (G) Association between Δage and fasting blood glucose (FBG). (H) Association between Δage and low-density lipoprotein (LDL) cholesterol.

**Supplement Figure 2.**
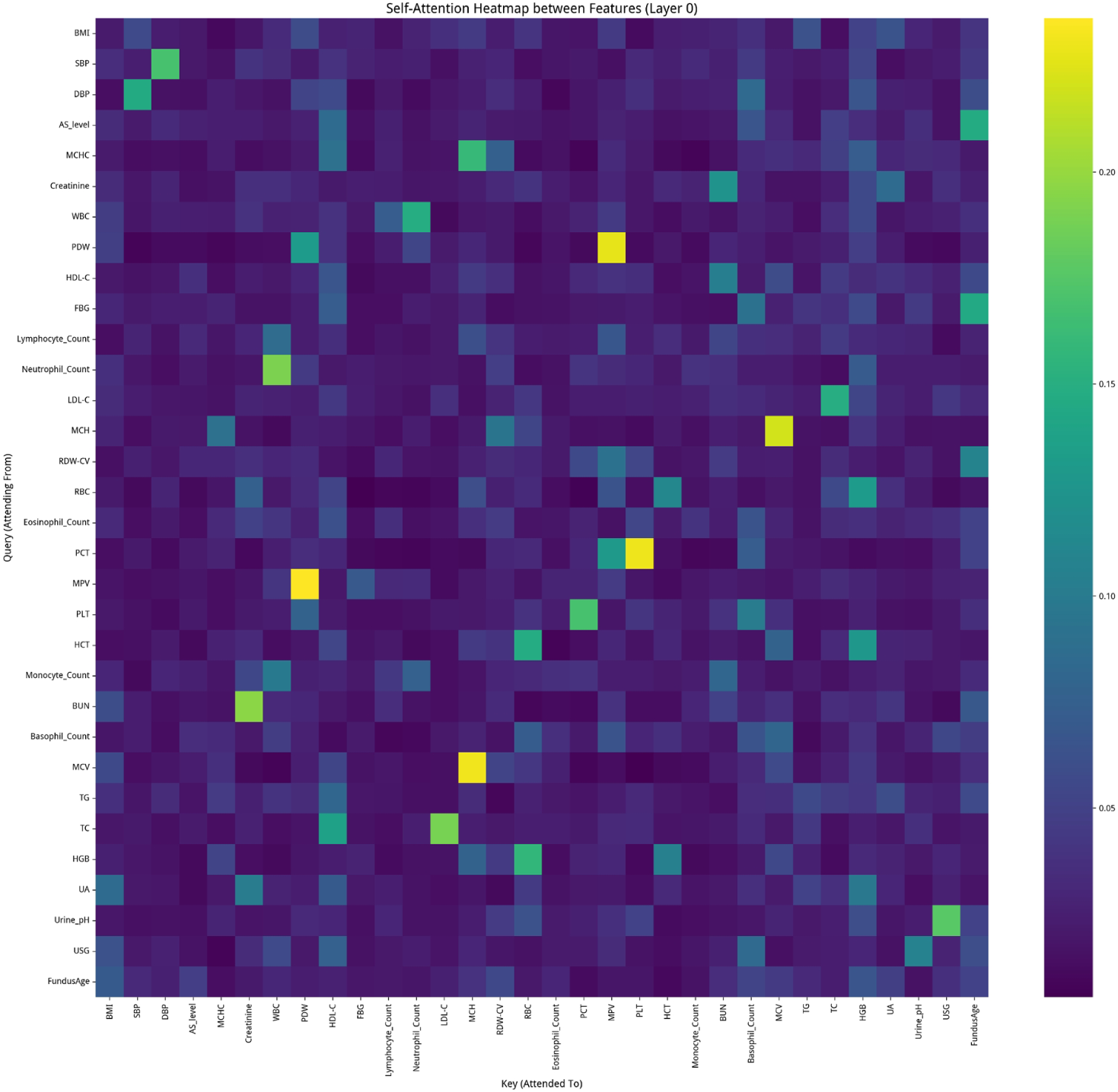
Attention heatmap from a Tabular Transformer analyzing associations between multi-dimensional indicators and predicted FundusAge. Attention heatmap from a Tabular Transformer analyzing associations between multi-dimensional indicators and predicted FundusAge. We constructed a Tabular Transformer, pretrained on data from 37,175 individuals (training set) and validated on 9,295 (test set), to generate this attention heatmap. The map illustrates internal information flow as the model builds feature representations (Y-axis: ‘query’ indicator; X-axis: ‘attended-to’ indicators). The model successfully captured clinically intuitive associations, as high-attention (bright) spots align with known biological correlations. In contrast, FundusAge exhibited low association with most other indicators, suggesting it is a novel and independent biomarker whose information cannot be easily explained by conventional clinical data.

**Supplement Figure 3.**
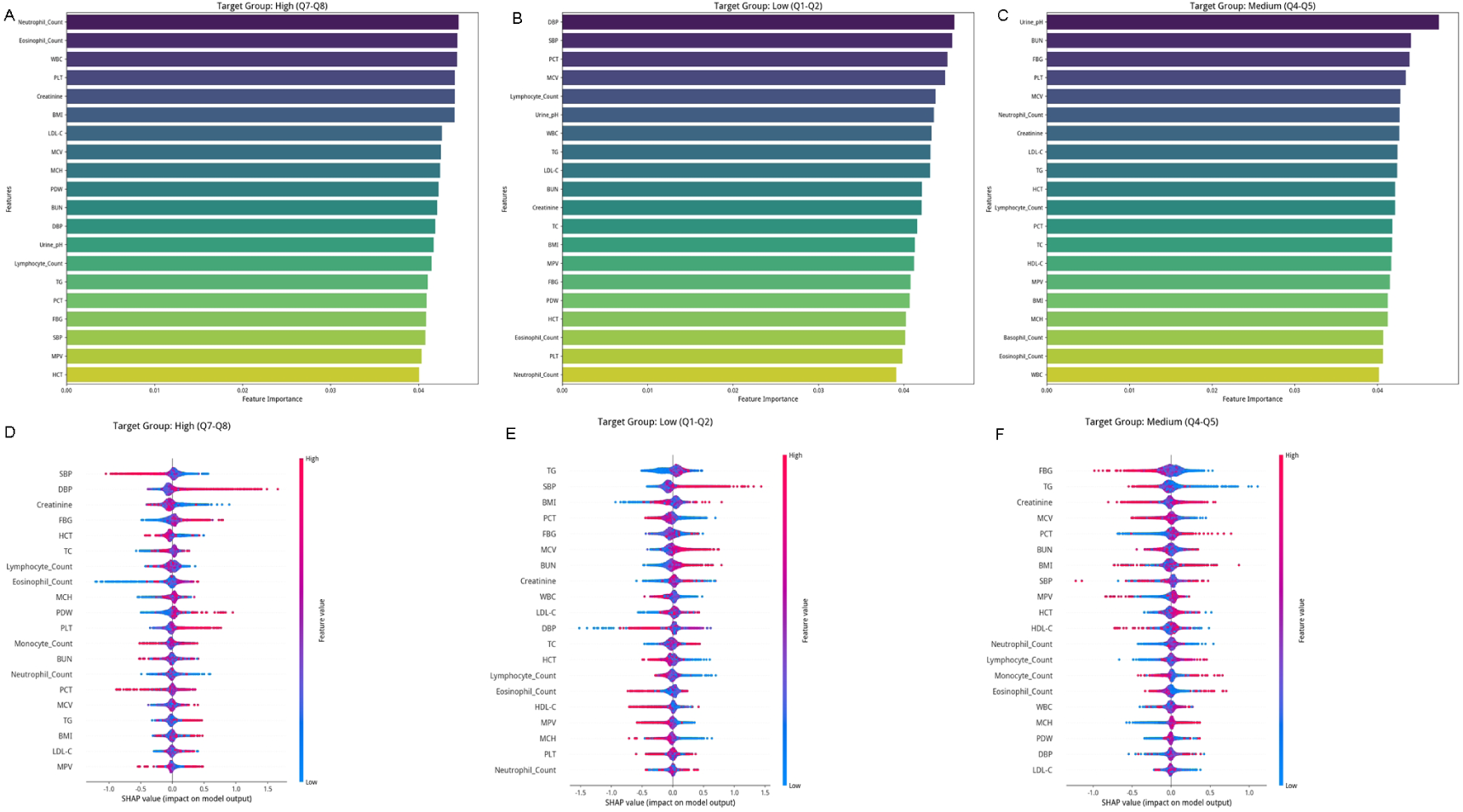
Analysis of XGBoost model results for classifying individuals into zones based on multi-dimensional physiological and biochemical indicators. We developed an XGBoost model to classify individuals into Low (Q1-Q2), Medium (Q4-Q5), or High (Q7-Q8) zones, achieving accuracies of 74.69% (Low), 74.72% (Medium), and 74.84% (High). The model evaluated the contribution of each input feature, with feature importance and SHAP beeswarm plots presented for each zone. (A-C) Feature importance plots of key drivers for the Q7-Q8 (A), Q1-Q2 (B), and Q4-Q5 (C) groups. (D-F) SHAP beeswarm plots of key drivers for the Q7-Q8 (D), Q1-Q2 (E), and Q4-Q5 (F) groups.

